# The effect of travel restrictions on the spread of the 2019 novel coronavirus (2019-nCoV) outbreak

**DOI:** 10.1101/2020.02.09.20021261

**Authors:** Matteo Chinazzi, Jessica T. Davis, Marco Ajelli, Corrado Gioannini, Maria Litvinova, Stefano Merler, Ana Pastore y Piontti, Luca Rossi, Kaiyuan Sun, Cécile Viboud, Xinyue Xiong, Hongjie Yu, M. Elizabeth Halloran, Ira M. Longini, Alessandro Vespignani

**Affiliations:** Laboratory for the Modeling of Biological and Socio-technical Systems, Northeastern University, Boston, MA USA; Bruno Kessler Foundation, Trento Italy; ISI Foundation, Turin, Italy; Fogarty International Center, NIH, USA; School of Public Health, Fudan University, Key Laboratory of Public Health Safety, Ministry of Education, Shanghai, China; Fred Hutchinson Cancer Research Center, Seattle, WA, USA; Department of Biostatistics, University of Washington, Seattle, WA. USA; Department of Biostatistics, College of Public Health and Health Professions, University of Florida, Gainesville, USA

## Abstract

Motivated by the rapid spread of a novel coronavirus (2019-nCoV) in Mainland China, we use a global metapopulation disease transmission model to project the impact of both domestic and international travel limitations on the national and international spread of the epidemic. The model is calibrated on the evidence of internationally imported cases before the implementation of the travel quarantine of Wuhan. By assuming a generation time of 7.5 days, the reproduction number is estimated to be 2.4 [90% CI 2.2-2.6]. The median estimate for number of cases before the travel ban implementation on January 23, 2020 is 58,956 [90% CI 40,759 - 87,471] in Wuhan and 3,491 [90% CI 1,924 - 7,360] in other locations in Mainland China. The model shows that as of January 23, most Chinese cities had already received a considerable number of infected cases, and the travel quarantine delays the overall epidemic progression by only 3 to 5 days. The travel quarantine has a more marked effect at the international scale, where we estimate the number of case importations to be reduced by 80% until the end of February. Modeling results also indicate that sustained 90% travel restrictions to and from Mainland China only modestly affect the epidemic trajectory unless combined with a 50% or higher reduction of transmission in the community.

---

Starting in December 2019, Chinese health authorities have been closely monitoring a cluster of pneumonia cases in the city of Wuhan, in Hubei province. It has been determined that the pathogen causing the viral pneumonia among affected individuals is a new coronavirus (2019-nCoV)(1). As of February 7th, 2020, a total of 31,213(2) cases have been detected and confirmed in Mainland China. Internationally, there are more than 200 additional cases detected and confirmed in 23 countries(3). In this work we model both the domestic and international spread of the 2019-nCoV epidemic. We estimate the effects of the travel ban implemented in Wuhan and the international travel restrictions adopted by a number of countries in early February, 2020.

To model the international spread of the 2019-nCoV outbreak we use the Global Epidemic and Mobility Model (GLEAM), an individual-based, stochastic, and spatial epidemic model (4; 5; 6; 7). GLEAM uses a metapopulation network approach integrated with real-world data, where the world is divided into sub-populations centered around major transportation hubs (usually airports). The subpopulations are connected by the flux of individuals traveling daily among them. The model includes over 3,200 sub-populations in roughly 200 different countries and territories. The airline transportation data consider daily origin-destination traffic flows from the Official Aviation Guide (OAG) and IATA databases (updated 2019), while ground mobility flows are derived by the analysis and modeling of data collected from the Offices of Statistics for 30 countries on 5 continents (5). Within each sub-population, the human-to-human transmission of 2019-nCoV is modeled using a compartmental representation of the disease where individuals can occupy one of the following compartments: Susceptible (*S*), Latent (*L*), Infectious (*I*) and Recovered (*R*). Susceptible individuals can acquire the virus through contacts with individuals in the infectious compartment, and become latent, meaning they are infected but can not transmit the disease yet. Latent individuals progress to the infectious stage with a rate inversely proportional to the latent period (which we assume to have the same duration as the incubation period), and infectious individuals progress into the removed stage with a rate inversely proportional to the infectious period. The sum of the mean latent and infectious periods defines the generation time. Removed individuals represent those who can no longer infect others, meaning they are recovered, isolated, hospitalized, or dead.

The model generates an ensemble of possible epidemic scenarios described by the number of newly generated infections, times of disease arrival in each subpopulation, and the number of traveling infection carriers. We assume a starting date of the epidemic that falls between 11/15/2019 and 12/1/2019, with 40 cases caused by zoonotic exposure(8; 9). The posterior distribution of the basic reproductive number *R*_0_ is estimated by exploring the likelihood of importation of 2019-nCoV cases to international locations. We have performed a sensitivity analysis considering different combinations of average latency and infectious periods, exploring a generation time (*T*_*g*_) interval ranging from 6 to 11 days based on plausible ranges from the SARS epidemic and recent analysis of the 2019-nCoV data (10; 11; 12; 13; 14; 15; 16). Details and sensitivity analysis are reported in the Supplementary Material. In the following we report the results for a generation time *T*_*g*_ =7.5 days (12). The obtained posterior distribution provides an average reproductive number *R*_0_= 2.4 [90% CI 2.2-2.6], and a doubling time measured at *T*_*d*_ = 4.6 days [90% CI 4.2-5.1]. The obtained values are in the same range as previous analyses based on early 2019-nCoV data (12; 17; 18; 19; 20). Although the calibration obtained for different generation times provides different posterior distributions for *R*_0_, the overall evolution of the epidemic is determined by the growth rate of infectious individuals and generates results consistent with those presented here. Results under the assumptions of a different generation time (*T*_*g*_ = 9), and the presence of mildly symptomatic individuals not detected internationally are reported in the Supplementary Material.

On January 22, 2020, the projected median number of cases with no travel restrictions for Mainland China excluding Wuhan is 3,491 [90% CI 1,924-7,360]. The overwhelming majority of cases are in Wuhan with a median number of 58,956 [90% CI 40,760-87,471]. To analyze the effect of the travel ban within Wuhan, we implemented long-range travel restrictions beginning on January 23rd and restricted the local commuting flows on January 25th. Initially, we assume no changes in the transmissibility and disease dynamics. The model output shows no noticeable differences in the epidemic trajectory of Wuhan, while it shows a delay of about 3 days occurring for other locations in China (see Fig. 1A). The overall reduction of cases in Mainland China excluding Wuhan is close to 20% by February 22, with a relative reduction of cases across specific locations varying in a range from 1% to 57%,(Fig. 2). With a doubling time of approximately 5 days, this level of reduction corresponds to only a modest delay of the epidemic trajectory of 1 to 6 days. These results are in agreement with estimates resulting from the combination of epidemiological and human mobility data (21). The model indicates clearly that as of January 23, 2020, the epidemic was seeded in several locations across Mainland China. As an independent validation test, we show in fig 1B) the cumulative number of cases in Mainland China provinces through February 5, 2020, as reported from DXY.cn, a Chinese online community network for physicians, health care professionals, pharmacies and healthcare facilities established in 2000 (22; 16), and compare these results with model projections. The model projections are highly correlated with the observed data (Pearson’s *r* = 0.77, *P <* 0.000001), although as expected we find that there are significantly fewer reported cases than projected (See Fig. 1 B). If we assume that the observed number of cases are the result of different binomial processes that with a certain probability will determine the actual detection of a case, we find that the median ascertainment rate of detecting an infected individual in the population is equal to 19.59% [IQR: 14.36%, 35.58%] in Mainland China. In other words, our model suggests that surveillance only detects one out of five cases.

**Figure 1:**
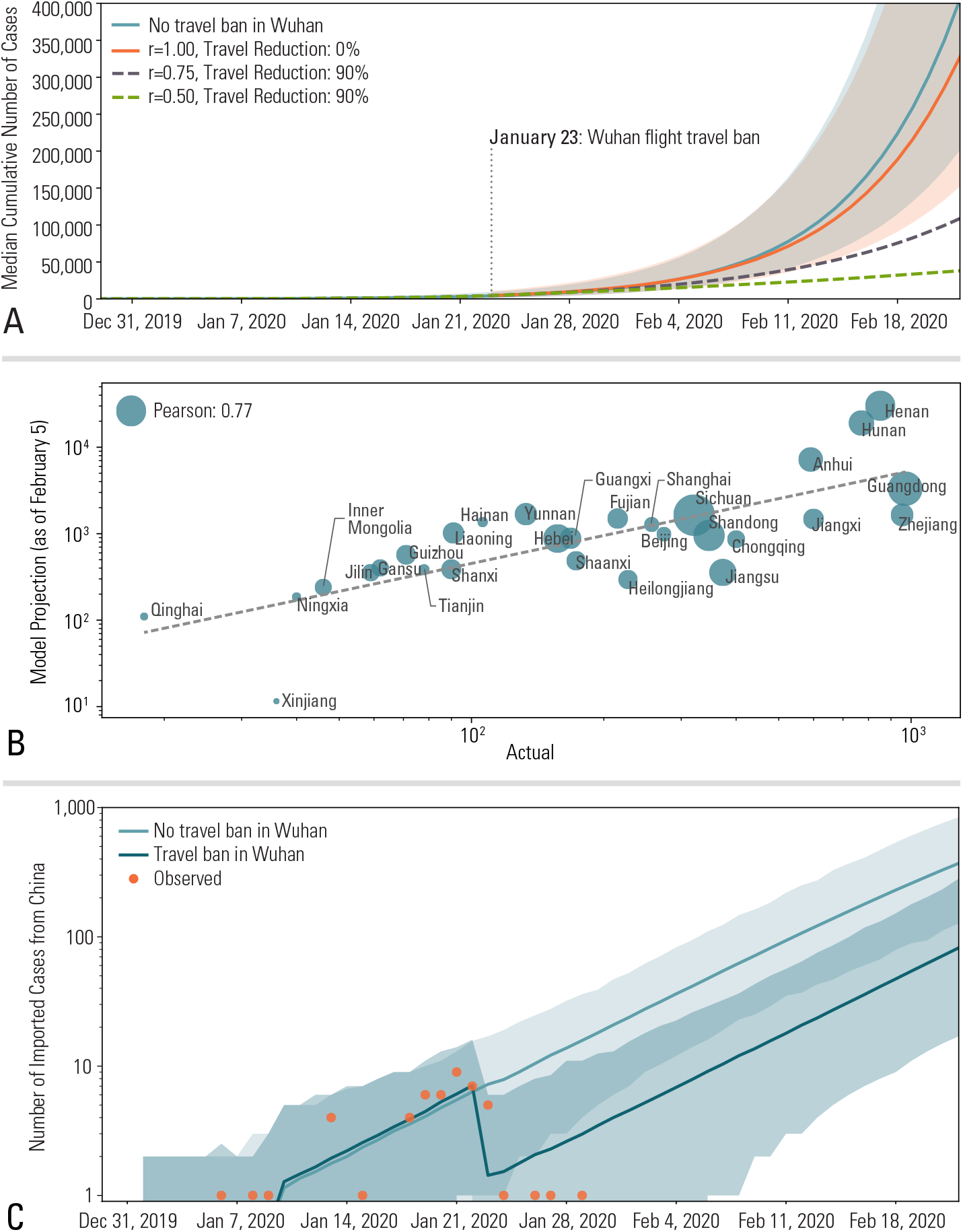
A) Trajectory of the 2019-nCoV epidemic in Chinese locations (excluding Wuhan) under the travel ban to and from Wuhan in effect as of January 23rd, 2020. The lines represent the median cumulative number of cases while the shaded areas represent the 90% reference range. B) Number of cases predicted by the model on February 5th as a function of the number cases observed in individual provinces in China by that date. The size of the circles are proportional to the population size in each province. C) Projections of the average total number (daily) of international case importations with and without travel ban from Wuhan. Observed data of international case importations with a travel history from Wuhan by arrival date. Shaded areas represent the 99% reference range.

**Figure 2:**
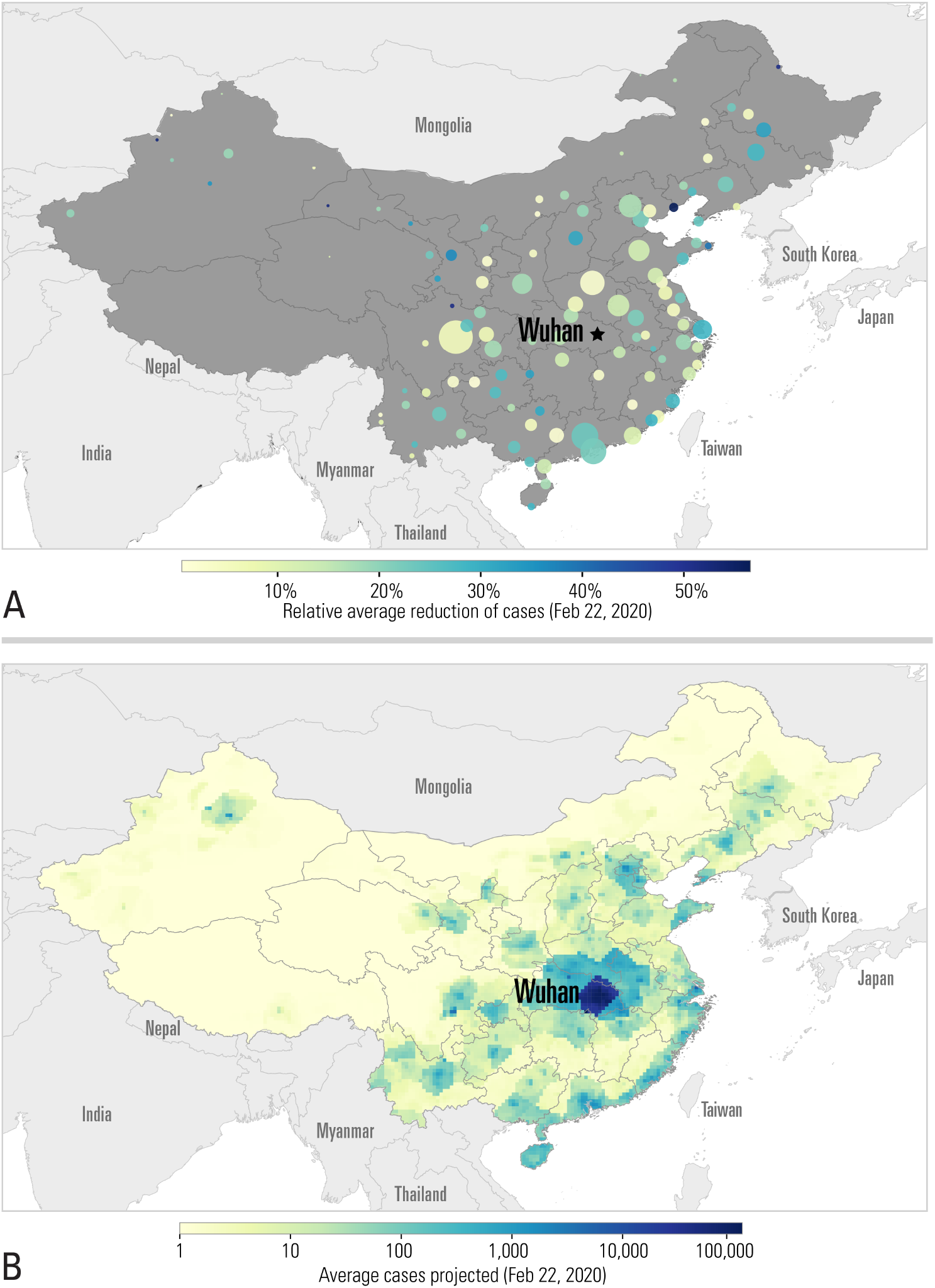
A) Relative reduction of incidence across China as of February 22, 2020. The color of circles represents the relative reduction in the number of cases, while the size represents the population in the region. B) Projected cumulative number of cases by the same date, after implementing travel restrictions in Wuhan.

The model allows us to estimate the number of case importations in international locations from Mainland China. We analyze how this number would increase according to projections in a fully status quo scenario compared to the presence of a travel ban. In Fig. 1C) we report the mean number of total international importation events in the two scenarios. We find an 80% reduction in cases imported from Mainland China to other countries until the end of February. While the number of cases imported internationally initially has a marked decrease, it picks up again in the following weeks with importation from other locations in China. The model indicates that after the travel restrictions in Wuhan are implemented on January 23, the top 5 ranked cities as the origin of international case importations are Shanghai, Beijing, Shenzhen, Guangzhou, and Kunming. Similarly, the model allows ranking countries across the world according to the relative risk of importing cases from Mainland China. More precisely the relative risk is defined for each country *Y*, as the relative probability *P* (*Y*) that a single infected individual travels from the index areas to that specific destination *Y*. In other words, given the occurrence of one exported case, *P* (*Y*) is the relative probability that the disease carrier will appear in location *Y*, with respect to any other possible location. This risk depends on the travel flow from cities in Mainland China to other countries and the disease prevalence in those cities. It is also worth remarking that our traffic flows are origin destination data that do not depend on traveling routes, and are a proxy for the actual mobility demand across cities. In Fig. 3 we provide a visualization of how each one of the top cities in China contribute to the relative risk of the top 20 countries at risk of importation for the dates before and after the travel ban in Wuhan was in effect. In particular, before the travel ban 84% of the internationally imported cases originated from Wuhan; while after the travel ban, the top 10 contributors to the relative risk are needed to account for at least ~ 80% of the internationally imported cases where the top three contributors are: Shanghai (27.85%), Beijing (14.28%), and Shenzhen (13.68%). In terms of relative risk of importation, the countries at higher risk of importation after the implementation of the Wuhan travel ban are: Japan (11.01% pre-travel ban, 13.97% post-travel ban), Thailand (22.89% pre-travel ban, 12.01% post-travel ban), Republic of Korea (7.48% pre-travel ban, 11.58% post-travel ban), Taiwan (9.32% pre-travel ban, 9.88% post-travel ban), and USA (4.66% pre-travel ban, 5.91% post-travel ban).

**Figure 3:**
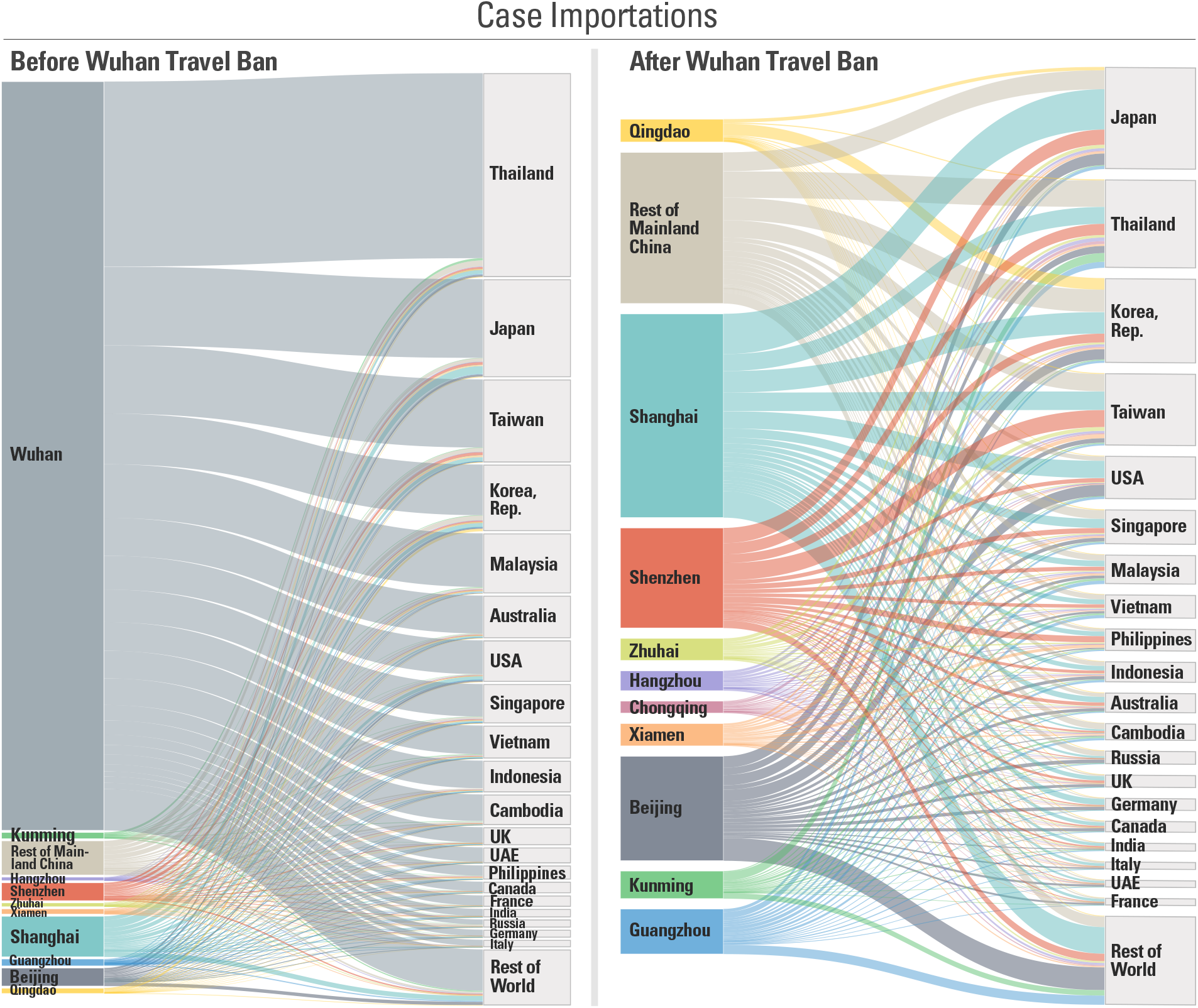
Contribution to the relative risk of importation of the top 10 Chinese cities (plus the rest of Mainland China) before and after the Wuhan travel ban. The listed countries correspond to the top 20 countries at risk of importation. Flows are proportional to the relative probability that any one imported case will be traveling from a destination to a target.

Starting early February 2020, 59 airline companies suspended or limited flights to Mainland China and a number of countries including USA, Russia, Australia, and Italy have also imposed government issued travel restrictions (23; 24; 25; 26; 27; 28). It is difficult to calculate exactly the level of traffic reduction imposed by these measures. For this reason, we analyze here two major scenarios in which travel restrictions produce a 40% and 90% overall traffic reduction to and from China. The same traffic reductions are also applied domestically in China. Along with travel reductions, we consider three scenarios concerning disease transmissibility: i) a status quo situation with the same transmissibility found from the model calibration through January 23, 2020; ii) a moderate relative reduction of the original transmissibility (25%), corresponding to a transmissibility dampening factor of *r* = 0.75; and iii) a high reduction (50%) of the original transmissibility (*r* = 0.50). This relative reduction of transmissibility could be achieved through early detection and isolation of cases, as well as behavioral changes and awareness of the disease in the population. In Fig. 4 we show the combined effects of the travel and transmissibility reductions on the epidemic incidence in Mainland China and the number of exported cases to other countries. The simulated scenarios show that even in the case of drastic travel reductions (Fig. 4D), if tranmissibility is not reduced (*r* = 1), the epidemic in China is delayed for no more than 2 weeks. The peak of the epidemic in Mainland China is reached at the end of April – early May, 2020. It is worth remarking that the epidemic peak in Wuhan in the absence of transmissibility reductions falls in the first week of March 2020. The number of cases imported to other countries, Fig. 4A-C, is initially affected by a tenfold reduction, but by March 1st 2020 when there is no transmissibility reduction (*r* = 1), this number has reached again the levels of 133 and 22 cases per day for the 40% and 90% travel restrictions scenarios, respectively. The concurrent presence of both travel and transmissibility reductions, however, produce a much larger synergistic effect visible by both delaying the epidemic activity in Mainland China and the number of imported cases. In the moderate transmissibility reduction scenarios (*r* = 0.75) the epidemic peak is delayed to late June 2020 and the total number of international case importations by March 1st 2020 are 21 and 3 cases per day for the 40% and 90% travel restrictions scenarios, respectively. Larger travel limitations (*>* 90%) will extend the period of time during which the importation of cases is greatly reduced. The relative reduction of 50% of the transmissibility (*r* = 0.5) along with travel restrictions does delay the epidemic growth in China that never surpasses the daily incidence of 1 case per 1,000 in Mainland China, and the number of imported cases at international destinations are always in the single digit range. The effect of the transmissibility reduction is visible also on the short term epidemic curve in Mainland China as shown in Fig. 1, with a drastic reduction of the growth of the number of cases by February 22, 2020 with respect to the status quo epidemic curve.

**Figure 4:**
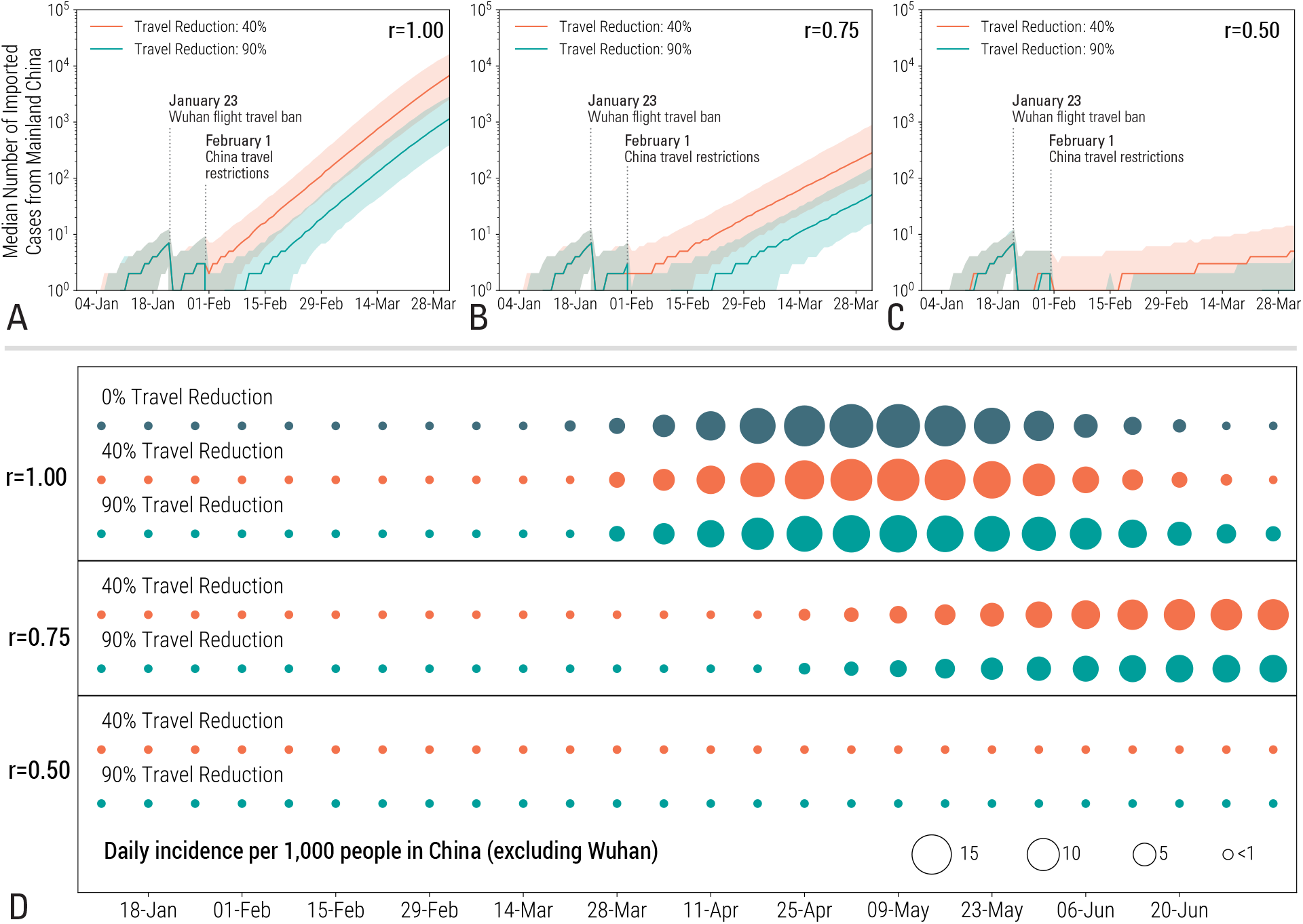
Analysis of the combined effects of travel and transmissibility reductions on the epidemic. Median number of imported cases from Mainland China for different dampening factors of the original transmissibility (r) and travel reductions (TR) A) no transmissability reduction, TR ∈{40%, 90%}; B) r = 0.75, TR∈ (40%, 90%; C) r = 0.5, TR∈{40%,90%} Shaded areas represent the 90% confidence interval. D) Incidence in Mainland China excluding Wuhan for the different scenarios considered in A-C

The presented analysis, as all modeling exercises, has a number of limitations and assumptions that are worth considering. The model parameters such as generation time and incubation period are chosen based on early results on the 2019-nCoV outbreak and prior knowledge of SARS and MERS coronavirus epidemiology. While we believe that the model is rather stable to variations in these parameters, more information on the key characteristic of the disease would considerably reduce uncertainties. The transmission and mobility model does not account, at this stage, for heterogeneities due to age differences in susceptibility and contact patterns. The calibration of the model does not consider correlations among importations (family travel) and assumes that travel probabilities are homogeneous across all individuals in the catchment area of each transportation hub. We were not able to find reliable data sources on the effectiveness of containment measures in place in Mainland China before Jan 23 (e.g. body temperature screening for departure in Wuhan International airport) which are thus not included in the model. In the travel restriction scenario we assume long term enforcement of individual mobility restrictions (travel was restricted until the end of June 2020). This might not be a feasible nor sustainable policy for such a long period of time.

Even in presence of the above limitations, the analysis of the Wuhan 2019-nCoV outbreak and the modeling assessment of the effects of travel limitations could be instrumental to national and international agencies for public health response planning. We show that by January 23 2020, the epidemic had already spread to other cities within China. The travel quarantine around Wuhan has only modestly delayed the epidemic spread to other areas of China. This is in agreement with separate studies on the diffusion of the 2019-nCoV virus in China (29; 21; 30). The model indicates that while the Wuhan travel ban was initially effective at reducing international case importations, the number of cases observed outside China will resume its growth after 2-3 weeks from cases that originated elsewhere. Furthermore, the modeling study shows that additional travel limitations up to 90% of the traffic have a modest effect unless paired with public health interventions and behavioral changes that achieve a considerable reduction in the disease transmissibility (31). The above results provides data with potential uses for the definition of optimized containment schemes and mitigation policies that includes the local and international dimension of the 2019-nCoV epidemic.

## Data Availability

The datasets generated and analysed during the current study are available from the corresponding author on reasonable request.

## Acknowledgements

MEH acknowledge the support of the MIDAS-U54GM111274. SM and MA acknowledge support from the EU H2020 MOOD project. CG and LR acknowledge support from the EU H2020 Icarus project. The findings and conclusions in this study are those of the authors and do not necessarily represent the official position of the funding agencies, the National Institutes of Health or U.S. Department of Health and Human Services.

## Notes

### Competing Interest Statement

All authors have completed the ICMJE uniform disclosure form at www.icmje.org/coi_disclosure.pdf and declare: no support from any organisation for the submitted work; MEH reports grants from National Institute of General Medical Sciences, during the conduct of the study; AV reports grants and personal fees from Metabiota inc., outside the submitted work; MC and APyP report grants from Metabiota inc., outside the submitted work; HY reports grants from Glaxosmithkline (China) Investment Co., Ltd, grants from Yichang HEC Changjiang Pharmaceutical Co., Ltd, grants from Sanofi Pasteur, grants from Shanghai Roche Pharmaceuticals Company, outside the submitted work. No other relationships or activities that could appear to have influenced the submitted work.

